# Anxiety and perceived risk during COVID-19 outbreak

**DOI:** 10.1101/2020.07.24.20161315

**Authors:** Chetna Malhotra, Isha Chaudhry, Semra Ozdemir, Irene Teo, Eric Andrew Finkelstein

**Author notes:** **Corresponding author**: Dr. Chetna Malhotra, Assistant Professor, Address: Duke-NUS Medical School, 8 College Road, Singapore 169857.

## Abstract

The uncertainty around coronavirus disease-19 (COVID-19) has triggered anxiety among public. We aimed to assess the variation in anxiety and risk perceptions of COVID-19 among adults in Singapore. We administered a web-survey to a panel of residents between 31 March and 14 April 2020. We assessed anxiety using general anxiety disorder (GAD) scale and assessed participants’ risk perceptions regarding severity of the outbreak. Of the 1,017 participants, 23% reported moderate to severe anxiety [GAD score≥10]. A high proportion reported perceived likelihood of ICU admission (46%) and death (30%) upon getting COVID-19. Results from path analysis showed that younger participants, those with chronic conditions, those living with children and low perceived trust in government response to COVID-19 had a significantly higher anxiety mediated by their perceived risk of dying upon getting COVID-19. These results highlight the need for management of anxiety through adequate and effective risk communication for the general public.

## Introduction

Coronavirus Disease-19 (COVID-19) is a major public health crisis affecting several nations (Dryhurst et al., 2020). This unprecedented infectious disease outbreak resulted in stringent public health measures around the word to curtail its spread (Qian et al., 2020).

In Singapore, the first confirmed case of COVID-19 was reported on January 23, 2020. By February 7, 2020, Singapore’s Disease Outbreak Response System Condition’ (DORSCON), a colour-coded framework for infectious disease outbreaks, turned orange, signifying that disease is severe and spreading easily. The uncertainty surrounding the COVID-19 outbreak including the lack of information regarding the risk of acquiring COVID-19, risk of developing complications and of dying due to it, and doubts about the adequacy of measures being taken to control the outbreak are likely to have created anxiety among people.

The primary objective of this study is thus to assess prevalence of anxiety among adults in Singapore during the COVID-19 outbreak. We hypothesize that older adults and those with pre-existing chronic medical conditions-groups at greater risk of experiencing adverse outcomes due to COVID-19 (Mueller, McNamara, & Sinclair, 2020) - will experience greater anxiety. We also hypothesize that people living with children and with lower trust in the government’s ability to control COVID-19 outbreak will have greater anxiety (Dryhurst et al., 2020; Shevlin et al., 2020). Lastly, we hypothesize that these associations will be mediated by perceived risk of: i) getting COVID-19 compared to a regular flu, ii) being admitted to Intensive Care Unit (ICU), and ii) death upon getting COVID-19.

## Methods

Between 31 March and 14 April 2020, we administered a web survey to a panel of residents (≥21 years) in Singapore. We quota sampled based on age, ethnicity and gender to ensure representativeness.

We assessed participants’ anxiety using general anxiety disorder scale (GAD-7) (Spitzer, Kroenke, Williams, & Löwe, 2006), and described their age, living arrangement (i.e. whether living with children), and chronic medical conditions (self-reported from a list of 15 conditions). Additionally, we assessed the extent of perceived trust in the government response to successfully contain COVID-19. We also assessed participants’ risk perceptions regarding the severity of COVID-19. First, participants reported their chances of getting COVID-19 compared to a regular flu. Response options included “much lower than flu”, “slightly lower than flu”, “same as flu”, “slightly higher than flu” or much higher than flu”. Additionally, respondents responded to the likelihood of ICU admission and likelihood of death due to COVID-19 on a 4-item Likert scale ranging from “very unlikely” to “very likely”.

We used path analysis to test our hypotheses. We estimated standardized coefficients (with 95% CI) and assessed the model fit (chi-square and p-value, root mean square error of approximation (RMSEA) (threshold <.05), and standardized root mean square residual (threshold <.05)).(Pituch & Stevens, 2015) We first assessed the significance of all hypothesized paths and then removed statistically insignificant (p ≥ 0.05) paths, and reassessed fit of the revised model. We used Stata 16.1 for analyses.

## Results

1,017 participants completed the survey. Over half the population were female (51%) and median (IQR) age was 40 years. Our sample included majority of the respondents with a university degree (58%) and nearly one-third of the respondents (34%) had a pre-existing chronic condition.

About a quarter (23%) of the respondents had symptoms of moderate and severe anxiety (GAD score≥10). Nearly a quarter (24%) felt that their chance of getting COVID-19 was lower than getting flu. Strikingly, a high proportion reported that they were likely to be admitted to an ICU (46%) and to die upon getting COVID-19 (30%) (Table 1).

**Table 1.** Sample characteristics, N=1017

|  | N (%) |
| --- | --- |
| Female | 519 (51.0) |
| Age |  |
| Median (IQR) | 40 (32, 52) |
| <30 years | 170 (16.7) |
| ≥30 - <65 years | 794 (78.1) |
| ≥65 years | 53 (5.2) |
| Ethnicity |  |
| Chinese | 815 (80.1) |
| Malay | 91 (9.0) |
| Indians and others | 111 (10.9) |
| Education |  |
| Below university<sup>±</sup> | 426 (41.9) |
| University or above | 591 (58.1) |
| Living with a child | 368 (36.2) |
| Number of chronic health conditions |  |
| None | 668 (65.7) |
| 1 | 168 (16.5) |
| 2 or more | 181 (17.8) |
| Types of chronic health conditions |  |
| High cholesterol | 145 (14.3) |
| High blood pressure | 119 (11.7) |
| Heart disease | 77 (7.6) |
| Diabetes | 52 (5.1) |
| Cancer | 18 (1.8) |
| Chronic kidney disease | 12 (1.2) |
| Other chronic conditions* | 165 (16.2) |
| Trust in government to successfully contain the spread of COVID-19 |  |
| Strongly agree/somewhat agree | 728 (71.6) |
| Neither agree/disagree, somewhat disagree or strongly disagree | 289 (28.4) |
| General Anxiety Disorder Score (range: 0 – 21) |  |
| Median (IQR) | 5 (1,9) |
| Mild anxiety (score: 5 – 9) | 289 (28.4) |
| Moderate anxiety (score: 10 – 14) | 167 (16.4) |
| Severe anxiety (score: ≥15) | 69 (6.8) |
| Measures of perceived risk |  |
| Chances of getting COVID-19 compared to regular flu |  |
| Much lower than flu | 91 (8.9) |
| Slightly lower than flu | 152 (14.9) |
| Same as flu | 314 (30.9) |
| Slightly higher than flu | 302 (29.7) |
| Much higher than flu | 158 (15.5) |
| Likelihood of being admitted to Intensive Care Unit upon getting COVID-19 |  |
| Very unlikely | 118 (11.6) |
|  | **N (%)** |
| Somewhat unlikely | 427 (42.0) |
| Somewhat likely | 318 (31.3) |
| Very likely | 154 (15.1) |
| **Likelihood of death upon getting to COVID-19** |  |
| Very unlikely | 249 (24.5) |
| Somewhat unlikely | 462 (45.4) |
| Somewhat likely | 237 (23.3) |
| Very likely | 69 (6.8) |
| <sup>±</sup>includes below primary, primary, secondary, vocational or diploma<br>*includes chronic- lung condition, gastro-intestinal disease, neurological condition, back pain, osteoporosis, joint-pain, arthritis, rheumatoid disease or nerve pain or eye problems |  |

Results from our final model (Figure 1) showed that those with – more chronic conditions, living with children and a lower perceived trust in government response to COVID-19 had significantly higher anxiety, which was mediated by their perceived risk of dying upon getting COVID-19. Contrary to our hypothesis, younger adults experienced higher anxiety, and this associated was not mediated by perceived risk of dying upon getting COVID-19. The model fit was good (chi-square (p-value) =4.043 (0.132); RMSEA=0.03; SMR=0.01). Perceived risk of getting COVID-19 compared to regular flu and risk of ICU admission upon getting COVID-19 did not mediate the association between any contextual factor and anxiety.

**Figure 1.**
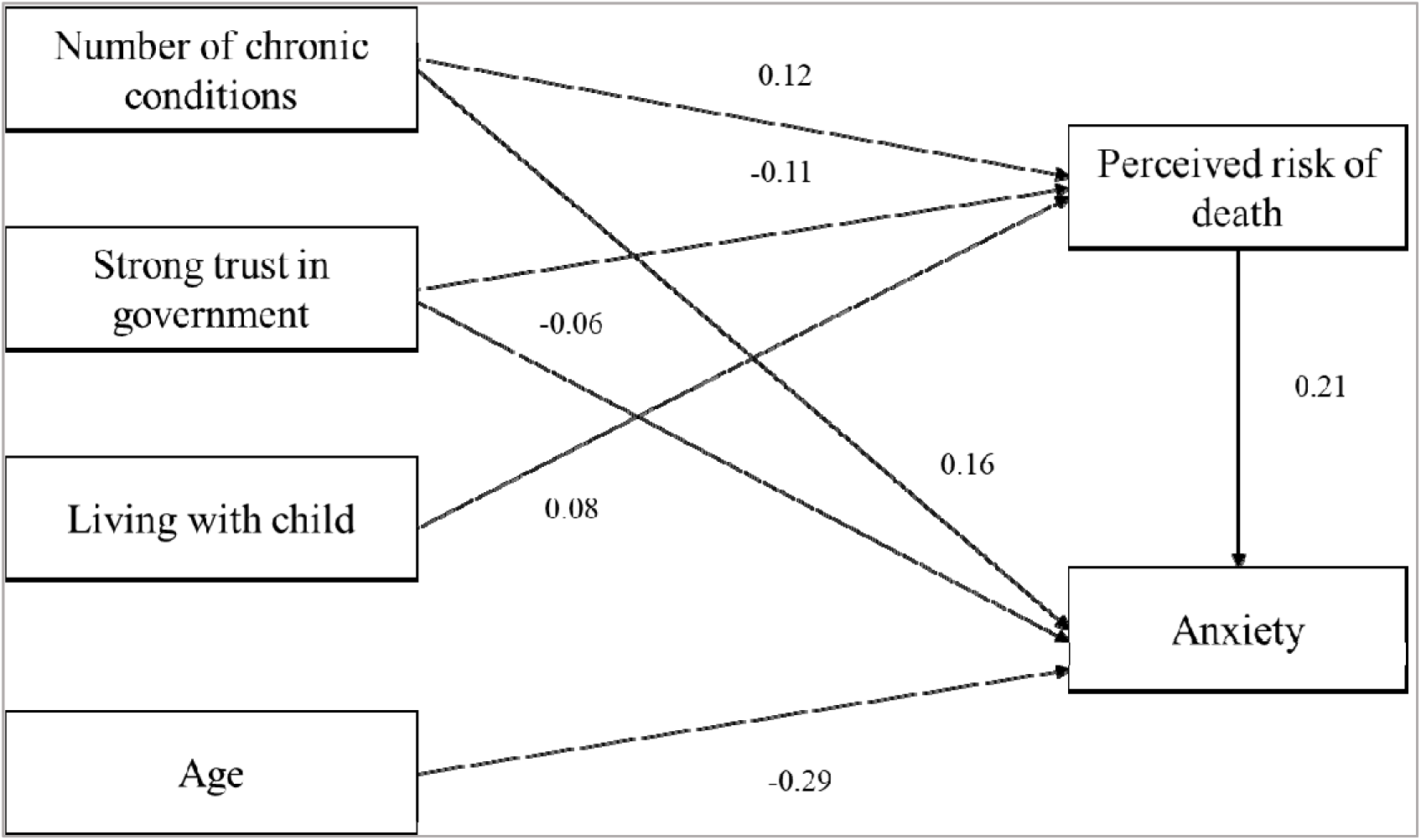
Path analysis of association between anxiety and perceived risk with standardized coefficients

## Discussion

A significant proportion of the participants reported moderate to severe anxiety (23%), much higher than what has been reported from population surveys before COVID-19 outbreak,(Lee, Sagayadevan, Vaingankar, Chong, & Subramaniam, 2015) and higher than those among health care professionals in Singapore (Tan et al., 2020). The high proportion of people who overestimated their risk of being admitted to ICU or dying if they get COVID-19 is also notable. As trust in government response to contain COVID-19 was high, we suggest that government agencies should carefully craft messages to communicate risk of adverse outcomes due to COVID-19 to the general public to help people clearly understand their risks and steps they can take to minimize this risk.

Future research can seek to better understand why younger adults are experiencing high anxiety. Since such high level of anxiety among the young population was not mediated by their perceived risk for getting COVID, being admitted to ICU or dying from COVID, it is possible that their anxiety is related to social isolation, loneliness, loss of income/employment opportunities or exposure to social media during COVID outbreak (Dryhurst et al., 2020). Population-level interventions (e.g. at educational institutes and workplaces) targeted at prevention and management of anxiety can help to reduce its serious or long-term sequel and are urgently required. Telehealth and online mental health consultations can be used to address the immediate mental health needs of the public.

A limitation of our study is that due to its cross-sectional design, we cannot infer causal relationship between perceived risk and anxiety. Although, we used a convenience sample of adults, our results concur with studies from other countries showing high population-level anxiety (Qian et al., 2020).

In conclusion, we find high anxiety among younger adults, those with chronic conditions, living with children and with low trust in the government response to control the outbreak. Effective communication should adequately convey the accurate information on risk of dying due to COVID-19 to address anxiety among public. Interventions to manage anxiety among younger adults are critical to improve their overall well-being during this and any future infectious disease outbreaks.

## Data Availability

The datasets generated and/or analysed during the current study can be made available from Dr. Malhotra as required.

## Source(s) of support/funding

This project is funded by Lien Centre for Palliative Care (LCPC Research N-911-000-030-091), Duke-NUS Medical School, Singapore

## Ethics^1^

National University of Singapore Institutional Review Board (Application Reference Number: S-20-085)

## Disclosure

Dr. Malhotra, Ms. Chaudhry, Dr. Ozdemir, Dr. Teo and Dr. Finkelstein have nothing to disclose.

## Footnotes

1 Study was exempted from review by the National University of Singapore Institutional Review Board as it involved a survey where participants were not identified, tracked, or at any risk.

## Notes

### Competing Interest Statement

The authors have declared no competing interest.

### Author Declarations

Study was exempted from review by the National University of Singapore Institutional Review Board as it involved a survey where participants were not identified, tracked, or at any risk. National University of Singapore Institutional Review Board (Application Reference Number: S-20-085)

